# SOCIAL MEDIA AS AN INFORMATION, EDUCATION, AND COMMUNICATION TOOL FOR RABIES PREVENTION: AN INTERVENTIONAL STUDY

**DOI:** 10.1101/2023.06.24.23291848

**Authors:** Jay Verma, Anurag Agarwal

## Abstract

**Introduction:** Rabies is a fatal disease that can be avoided by treating animal bites promptly. Hence, post-exposure prophylaxis is critical. As a result, the National Rabies Control Program was approved under the 12th five-year plan. One of its strategies is to engage in Information, Education and Communication activities. Social media provides an opportunity for the quick and easy dissemination of research but is constrained by a lack of peer review and the risk of misinterpretation. The efficacy of a novel social media-based knowledge dissemination strategy for rabies prevention was tested in this study.

**Methods:** An experimental study design was followed, wherein 144 preclinical medical students were included in each control and test group. The test group was administered the intervention, which exposed the participants to health education material via social media across a span of 30 days. Participants’ knowledge, attitude, and practices were observed before and after the study duration.

**Results:** There was no significant change in the responses of the control (chi-square statistic = 2.46, p-value = 0.96) and test (chi-square statistic = 1.64, p-value = 0.98) groups across the study duration. A comparison of the change in responses between the two groups revealed an insignificant difference (chi-square statistic = 1.29, p-value = 0.73).

**Conclusion:** Social media is an ineffective strategy for improvements in health literacy over a short duration. Such strategies need further validation for longer durations. Moreover, the effectiveness of the health education material itself needs to be assessed.

## INTRODUCTION

Rabies is an invariably fatal viral disease that contributes to considerable morbidity and mortality throughout the Indian mainland. The virus is found in both domestic and wild animals but dog bites are responsible for most human transmission in India. Since rabies can be prevented by timely management of the animal bite, post-exposure prophylaxis (PEP) assumes much importance.^1,2^ So, the National Rabies Control Program (NRCP) was approved under the 12th five-year plan, consisting of both human and animal components.^1,2^ One of its strategies includes Information, Education, and Communication (IEC) activities, a public health approach aimed at educating the community about animal bite management, wound washing, and driving appropriate health-seeking behavior. This material is translated into local languages and disseminated as hoardings, radio fillers, and posters in schools and clinics.^1,2^

Traditionally, researchers have used methods like conferences and journal articles to share their work. While appropriate for the professional population, these methods have limited and differential reaches for the public and mass media. However, social media (“online platforms that allow users to generate content, exchange information, and communicate with one another”) has the potential to allow the effective dissemination of health information.^3^ After the COVID-19 social media “infodemic” emerged, significant research has been directed toward uncovering the advantages and limitations of social media for translational medicine. It presents great opportunities for rapid and easy dissemination of research, networking, and big data analytics. Still, it is limited by a lack of peer review and the risks of misinformation spread and misinterpretation on a large scale.^3-5^ As a result, the formulation of optimal practices to employ social media in the packaging and dissemination of health research is also being done.^6^

Further work is still needed to evaluate the role of social media in health research in the context of the Indian population since factors affecting such outcomes are population-specific. Considering the goals of the IEC system, it has much to gain from tapping into its potential benefits. A literature review revealed that such practices hadn’t been used for IEC material created under the aegis of the NRCP. The goal of this study is to evaluate the effects of social media on information dissemination and retention for NRCP IEC material (specifically, among preclinical medical students) and provide evidence-based recommendations for using such strategies in the best possible manner.

## METHODS

### Study Design

An interventional study design was followed. Individuals in the sampled population were randomly divided into two groups: those exposed to the intervention (test group) and those not exposed to the intervention (control group). The study was divided into 3 phases: pre-intervention, intervention, and post-intervention. During the 1st phase, baseline data was collected. During the 2nd phase, participants were exposed to the intervention for 30 days. The 3rd phase began 15 days after the 2nd phase ended and the same data collection methods were used to measure the impact of the intervention on information dissemination and retention, followed by data analysis.

### Intervention

The test group was administered the rabies IEC material available on the National Center for Disease Control (NCDC) website (https://ncdc.gov.in/index1.php?page=1&ipp=All&lang=1&level=2&sublinkid=502&lid=428) in English and Hindi languages using a WhatsApp broadcast every 3 days for 30 days, in a cyclical manner. Text messages, encouraging the participants to go through the material, were also a part of this intervention.

Even though it’s debated whether WhatsApp falls under the definition of “social media”, it allows users to create content (up to a limited extent) and share it, thus having certain features of social media. Its usage in this study is justified by its popularity in the study population as a messaging app.

### Study Population

The study population included preclinical medical students, i.e., Bachelor of Medicine, Bachelor of Surgery (MBBS) first and second-professional-year students who have a smartphone, internet access, and are WhatsApp users. Individuals who themselves had been dog bite or rabies victims or had a dog bite or rabies victim in the household were excluded. Individuals who developed the aforementioned exclusion criteria during the study duration were excluded from the data analysis. Due to the short study duration of 2 months, only preclinical medical students of Maulana Azad Medical College, Delhi, India were sampled.

### Sampling Procedure

Simple random sampling was used to select potential participants. Since the sample size was calculated to be less than 10% of the population size, sampling without replacement was deemed to be appropriate.

### Sample Size

The sample size was calculated using the open-source software OpenEpi (https://www.openepi.com/Menu/OE_Menu.htm). Setting a two-sided confidence interval as 95%, the power as 80%, the ratio of unexposed to exposed individuals in the sample as 1, a statistically significant magnitude of the expected effect (prevalence/risk ratio) as 3, and assuming the percentage of the unexposed population to have desired outcomes as 5% (due to a lack of previous records on this parameter), a sample size of 142 individuals each for test and control groups were obtained using Kelsey’s formula.

### Data Collection

The questionnaire (Appendix A) was designed to assess the knowledge, attitude, and practices of the study participants about animal bite management, wound washing, and healthcare-seeking behavior after a dog bite. The primary outcome is a change in awareness about animal bite management and the secondary outcomes include a change in awareness about rabies transmission and healthcare-seeking behavior after an animal bite. It was administered two times: pre-intervention and post-intervention phases. Since the questionnaire is not pre-validated, it was tested on 20 participants as well as institutional scientific committee approval was obtained before data collection.

### Data Analysis

The responses obtained through the KAP questionnaire were entered into an MS Excel file and analyzed using SPSS version 24. The responses were scored +1 for the correct answer, -1 for the incorrect answer, and 0 for the “Don’t know” answer. Aggregated scores were categorized using Bloom’s cut-off points for each of the 3 sections (knowledge, attitude, and practices): “good”, if the score was between 80 and 100%, “moderate” if the score was between 60 and 79%, and “poor” if the score was less than 60%. Means and percentages of responses were calculated for both test and control groups, before and after the intervention. The changes in responses for both groups were measured and compared. Any association that may exist between the change in responses and being a dog/cat owner was also investigated. Hypothesis testing was done using Chi-square tests and a p-value of less than or equal to 0.05 was considered statistically significant. Data analysis for the final qualitative question was done by categorizing the responses and calculating the percentage of each category.

### Ethical Considerations

Due to the involvement of human subjects, Institutional Ethics Committee approval was obtained before the study began (Institutional Ethics Committee Protocol Identity: F.1/IEC/MAMC/90/02/2022/No.112). The study has also been registered on *clinicaltrials.gov* (Identifier: NCT05702008). An informed consent form (Appendix B) was administered to potential participants with due diligence.

## RESULTS

### Participant Responses: Pre-intervention Phase

A total of 288 participants were enrolled in the study. Participants were randomly assigned to either the test or control group after their enrolment in the study and responses were collected during the pre-intervention phase to establish the baseline characteristics of the study population. They have been summarized in **Table 1**.

**Table 1.** Pre- and Post-Intervention Responses The table summarizes the pre- and post-intervention responses (aggregate scores) from the test group and the control group for each of the four domains in the questionnaire; aggregate scores are accompanied by the mean scores and Bloom’s cut-off points in parentheses.

|  | Knowledge of Rabies | Knowledge of Rabies Post- | Attitude Towards Dogs | Dog Bite Management |
| --- | --- | --- | --- | --- |

Table 1: The table summarizes the pre- and post-intervention responses (aggregate scores) from the test group 3 and the control group for each of the four domains in the questionnaire; aggregate scores are accompanied by 4 the mean scores and Bloom's cut-off points in parentheses.
|  | Transmission | exposure<br>Prophylaxis | and Dog Bites | Practices |
| --- | --- | --- | --- | --- |
| *Control Group* |  |  |  |  |
| Pre-intervention<br>Responses | 881/1440<br>(0.61,<br>moderate) | -140/1440 (-<br>0.09, poor) | 735/1440<br>(0.51, poor) | 675/1152<br>(0.58, poor) |
| Post-intervention<br>Responses | 913/1440<br>(0.63,<br>moderate) | -121/1440 (-<br>0.08, poor) | 795/1440<br>(0.55, poor) | 720/1152<br>(0.62,<br>moderate) |
| *Test Group* |  |  |  |  |
| Pre-intervention<br>Responses | 877/1440<br>(0.60,<br>moderate) | -127/1440 (-<br>0.08, poor) | 771/1440<br>(0.53, poor) | 711/1152<br>(0.61,<br>moderate) |
| Post-intervention<br>Responses | 914/1440<br>(0.63,<br>moderate) | -113/1440 (-<br>0.07, poor) | 838/1440<br>(0.58,<br>moderate) | 759/1152<br>(0.65,<br>moderate) |

### Participant Responses: Post-intervention Phase

The intervention (as described under Materials and Methods) was administered to the participants for 30 days and post-intervention data was collected using the same questionnaire as in the pre-intervention phase 15 days after the interventional phase ended. The responses have been summarized in **Table 1**.

For the final optional qualitative question in the questionnaire, participants did not give a significant number of responses for the investigator to make any meaningful conclusions. Therefore, rigorous data analysis for the final question has been omitted. The handful of responses that were obtained centered upon a desire for increased clarity in the IEC material on rabies immunoglobulin, the rabies vaccine, and their dosing regimens.

### Change in Responses

To evaluate whether there was any significant change in the responses, the chi-square test for categorical data was used. These results have been summarized in **Tables 2**.

**Table 2.** Change in Responses Across the Study Duration The table summarizes the responses from the control and test groups for each of the four domains in the questionnaire; aggregate scores are accompanied by the mean scores and Bloom’s cut-off points in parentheses; significance level = 0.05; chi-square statistic = 2.46977 (control group), 1.64875 (test group); p-value = 0.96 (control group), 0.98 (test group).

|  | Knowledge of<br>Rabies<br>Transmission | Knowledge of<br>Rabies Post-exposure<br>Prophylaxis | Attitude<br>Towards Dogs<br>and Dog Bites | Dog Bite<br>Management<br>Practices |
| --- | --- | --- | --- | --- |
| *Control Group* |  |  |  |  |
| Pre-intervention<br>Responses | 881/1440<br>(0.61,<br>moderate) | -140/1440 (-<br>0.09, poor) | 735/1440<br>(0.51, poor) | 675/1152<br>(0.58, poor) |
| Post-intervention Responses | 913/1440<br>(0.63, moderate) | -121/1440 (-0.08, poor) | 795/1440<br>(0.55, poor) | 720/1152<br>(0.62, moderate) |
| *Test Group* |  |  |  |  |
| Pre-intervention Responses | 877/1440<br>(0.60, moderate) | -127/1440 (-0.08, poor) | 771/1440<br>(0.53, poor) | 711/1152<br>(0.61, moderate) |
| Post-intervention Responses | 914/1440<br>(0.63, moderate) | -113/1440 (-0.07, poor) | 838/1440<br>(0.58, moderate) | 759/1152<br>(0.65, moderate) |

As is evident from the high p-values obtained from the data presented in tables 3 and 4, there was no significant change within the control and test group responses across the study duration.

**Table 3.** Comparison of the Change in Responses Across the Study Duration The table summarizes the change in aggregate responses across the study duration from the test group and the control group for each of the four domains in the questionnaire, accompanied by the percentage change and the change in mean responses in, parentheses; significance level = 0.05; chi-square statistic = 1.2932; p-value = 0.73.

|  | Knowledge of Rabies Transmission | Knowledge of Rabies Post-exposure Prophylaxis | Attitude Towards Dogs and Dog Bites | Dog Bite Management Practices |
| --- | --- | --- | --- | --- |
| Control Group | 32 (2.22%, 0.02) | 19 (1.32%, 0.01) | 60 (4.16%, 0.04) | 45 (3.90%, 0.04) |
| Test Group | 37 (2.56%, 0.03) | 14 (0.97%, 0.01) | 67 (4.65%, 0.05) | 48 (4.16%, 0.04) |

### Comparison of the Change in Responses

Across the study duration, the differences in responses were calculated to evaluate the efficacy of the social media-based intervention as an information dissemination strategy. These changes have been summarized in **Table 3**.

Based on the change in responses across the study duration, the value of the chi-square statistic was found to be 1.2932, which corresponds to a p-value of 0.73. Since the significance level was set to 0.05, the results are not statistically significant.

## DISCUSSION

In this study, the efficacy of a social media-based knowledge dissemination strategy has been tested, specifically for rabies prevention. The results show a very marginal improvement in the knowledge, attitude, and practices of rabies prevention, although the improvement in responses does not reach statistical significance. This suggests that social media cannot be a useful tool for rabies prevention and healthcare knowledge translation for short durations, in general.

The chief strength of this study was the randomized, controlled study design, of which there has been a lack in the field of health literacy. However, certain limitations associated with the interpretation of the results must be acknowledged. Of these, the primary one is the narrow nature of the study population which limits the applicability of the results to the general public; as a result, implications for policy-making and healthcare programs are greatly affected. Since medical students are already somewhat apprised of the rabies virus and its disease, it seriously confounds the extrapolation of these results toward the lay population. Moreover, due to scanty existing data on the tested parameters, the investigator had to rely on certain statistical assumptions to calculate the sample size. Another factor that limited the efficacy of the intervention was the short study duration of 2 months. As previous research shows, such interventions should be intense and focused over a longer period. Therefore, for policy-makers and public health professionals, it is of vital importance to assess the effectiveness of social media-based methods over years, when including them in healthcare programs.

Having considered the above limitations, the findings of this study might be called preliminary. Nevertheless, the results cannot be dismissed based on the limitations alone. In line with previous literature, they somewhat indicate the usefulness of social media as an information dissemination tool. Upon analyzing the results, it is evident that the IEC material formulated under the NRCP falls short concerning the messages it was intended to distill in the domain of rabies immunoglobulin knowledge, as shown by the relatively poor performance score in this domain. Given the complexity of this domain, the NRCP can certainly benefit from more work on simplifying this material so that even the general public may optimize their behavior regarding rabies immunoglobulin administration. The healthcare system may gain a lot in terms of reduced confusion among patients about rabies immunoglobulin, its benefits, and its limitations.

A comparison with previous research shows that social networking sites have been explored as health communication tools, albeit not in the Indian healthcare context. Greene, Choudhry, et al., 2010 evaluated the content of diabetes communication groups on Facebook, qualitatively.^7^ It showed that patients indeed tend to look for health advice on the Internet. Similarly, Liang and Scammon, 2011 provided a more quantitative analysis of social networking content on obesity.^8^ They provided knowledge on the support-seeking and support-giving behavior of patients. It follows from their findings that patients would benefit from the creation of a single, validated resource as it would reassure them and decrease the confusion centered around their treatments. While these studies provide an observational perspective on the impact of social networking sites, they do not have a randomized, controlled study design to demonstrate their efficacy. To provide a more comparative perspective, Rajagopalan, Khanna, et al., 2011 showed that non-professionally-maintained resources, even when they are of similar depth and accuracy as that of professionally-maintained resources, are less readable and hence, do not well-serve their purpose of health communication.^9^ Based on this, it can be commented that the creation of IEC material under the NRCP is more useful than invalidated resources, although, in absolute terms, more readability and clarity were desirable, as evaluated by the final question presented in the study questionnaire. Nordqvist, Hanberger, et al., 2009 showed that even healthcare professionals reacted positively to the use of social media for health communication.^10^

Particular attention is needed into uncovering the sociocultural and economic factors that lead to the development of inappropriate rabies prevention behavior. Since these factors tend to be population-specific, these researches should be targeted toward certain healthcare systems or geographic regions. Once that is done, tackling them by using the existing NRCP strategies and integrating them with social media-based methods is likely to make the whole program more efficient. Furthermore, evidence-based techniques like behavior change models should also be tested and employed. In parallel to improvement in the health literacy aspects, strengthening laboratory and vaccination aspects would also be required to sustain the changes. An important area of inquiry would be to shed light upon the most effective combinations of knowledge dissemination techniques such as text messages, short video films, posters, etc.

## CONCLUSION

Rabies is a fatal viral disease that causes significant morbidity and mortality across the Indian subcontinent. As a result, the National Rabies Control Program was approved under the 12th five-year plan. One of it strategies is to engage in Information, Education, and Communication activities, which is a public health approach aimed at educating the community about animal bite management, wound washing, and encouraging appropriate health-seeking behavior. In this study, the efficacy of a social media as a knowledge dissemination tool for rabies prevention was explored. The results show an improvement in rabies prevention knowledge, attitude, and practices, though the improvement in responses does not reach statistical significance. This suggests that social media might be a useful tool for rabies prevention and, more broadly, healthcare knowledge translation, but more thorough research methodologies are required.

## Data Availability

All data produced in the present study are available upon reasonable request to the authors.

## Conflict of interest statement by authors

The authors declare no conflict of interest for this research.

## APPENDICES

### Appendix A

Study Questionnaire

#### QUESTIONNAIRE

##### Date

###### Part I: Participant Information

1. Roll number:
2. Contact number:

###### Part II: Knowledge of Rabies Transmission

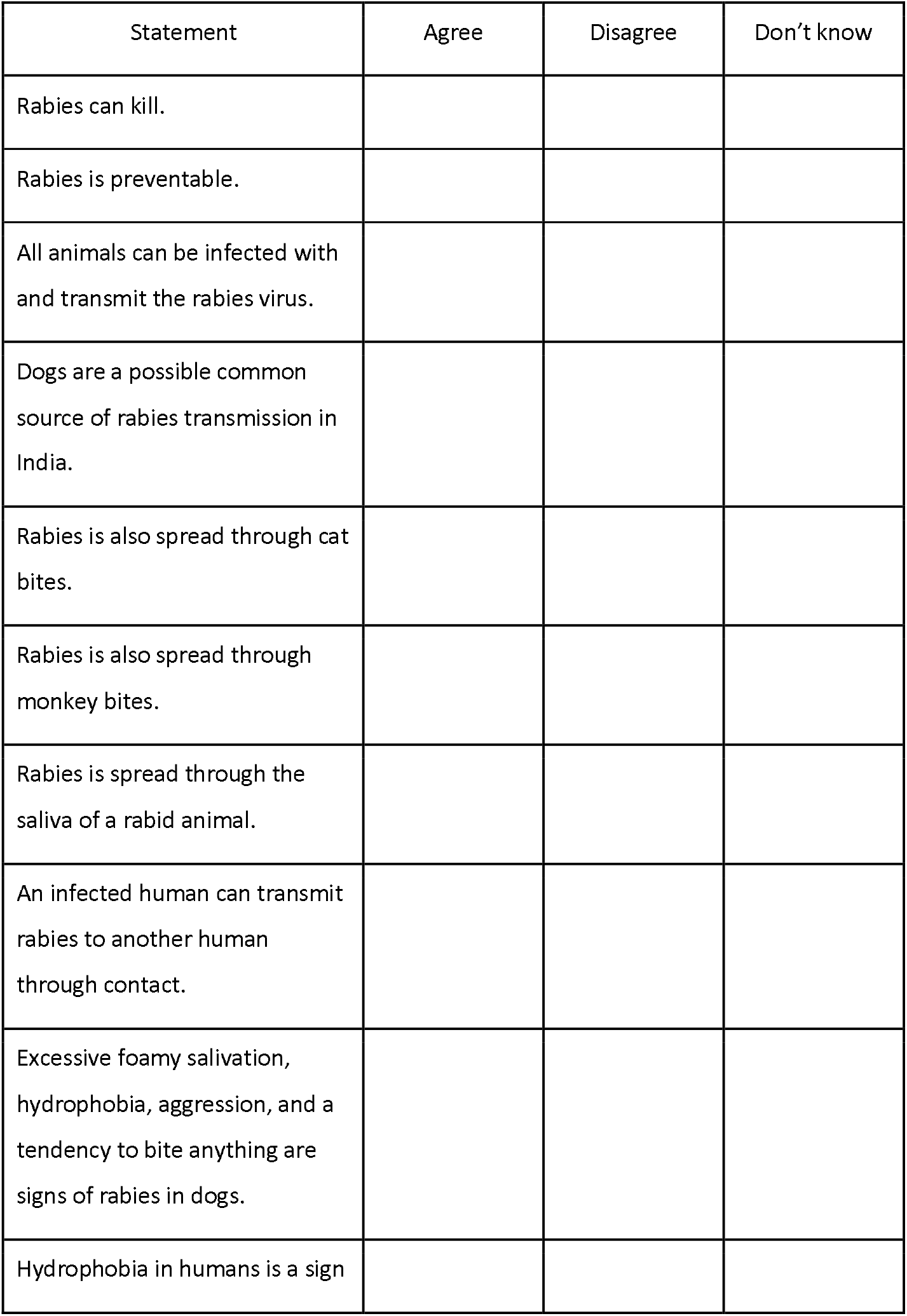
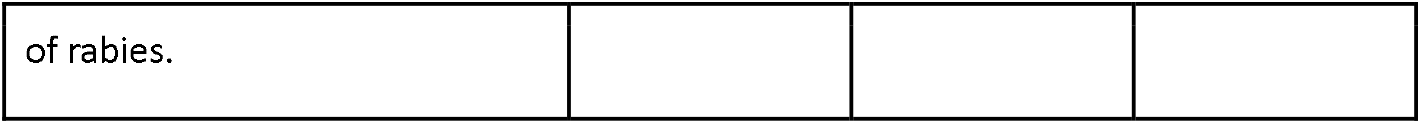

###### Part III: Knowledge of Rabies Post-Exposure Prophylaxis

| Statement | Agree | Disagree | Don't know |
| --- | --- | --- | --- |
| The first dose of Rabies Immunoglobulin (RIG) should be administered the same day as the dog bite. |  |  |  |
| RIG can be administered through intramuscular or intradermal routes. |  |  |  |
| For adults, RIG is administered in the deltoid region through the intramuscular route. |  |  |  |
| For infants and small children, RIG is administered in the anterolateral thigh region through the intramuscular route. |  |  |  |
| RIG can also be administered in the gluteal region through the intramuscular route (for both children and adults). |  |  |  |
| The intramuscular regimen requires a total of 5 visits to the doctor for RIG administration. |  |  |  |
| RIG is administered in the deltoid region on both sides through the intradermal route. |  |  |  |
| The intradermal regimen requires a total of 4 visits to the doctor for RIG administration. |  |  |  |
| For immunocompromised individuals, only the intramuscular route is recommended for RIG administration. |  |  |  |
| The National Rabies Control Program recommends the intradermal route for RIG administration. |  |  |  |

###### Part IV: Attitude Towards Dogs and Dog Bites

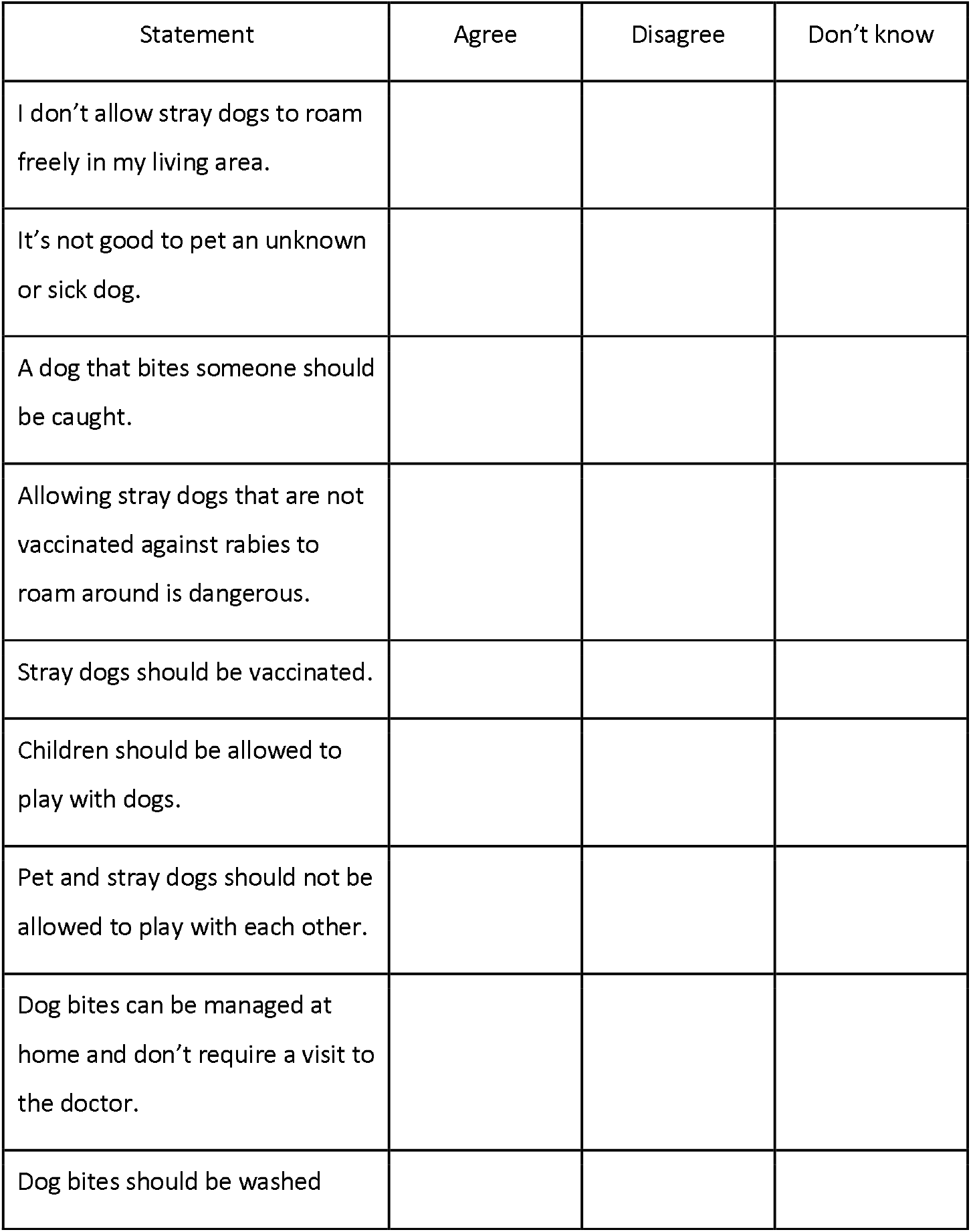
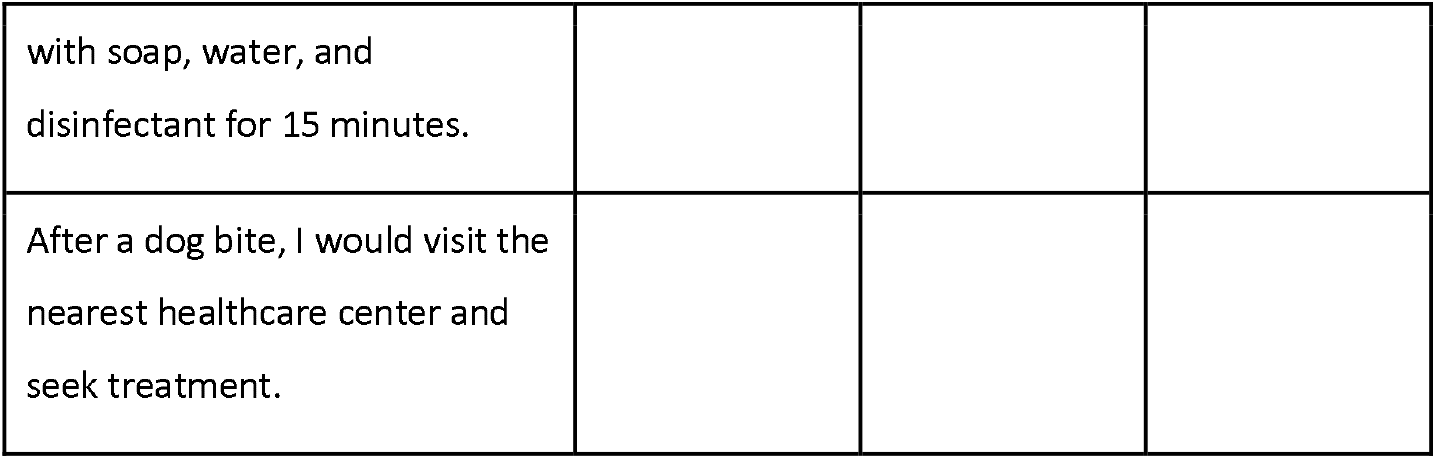

###### Part V: Dog Bite Management Practices

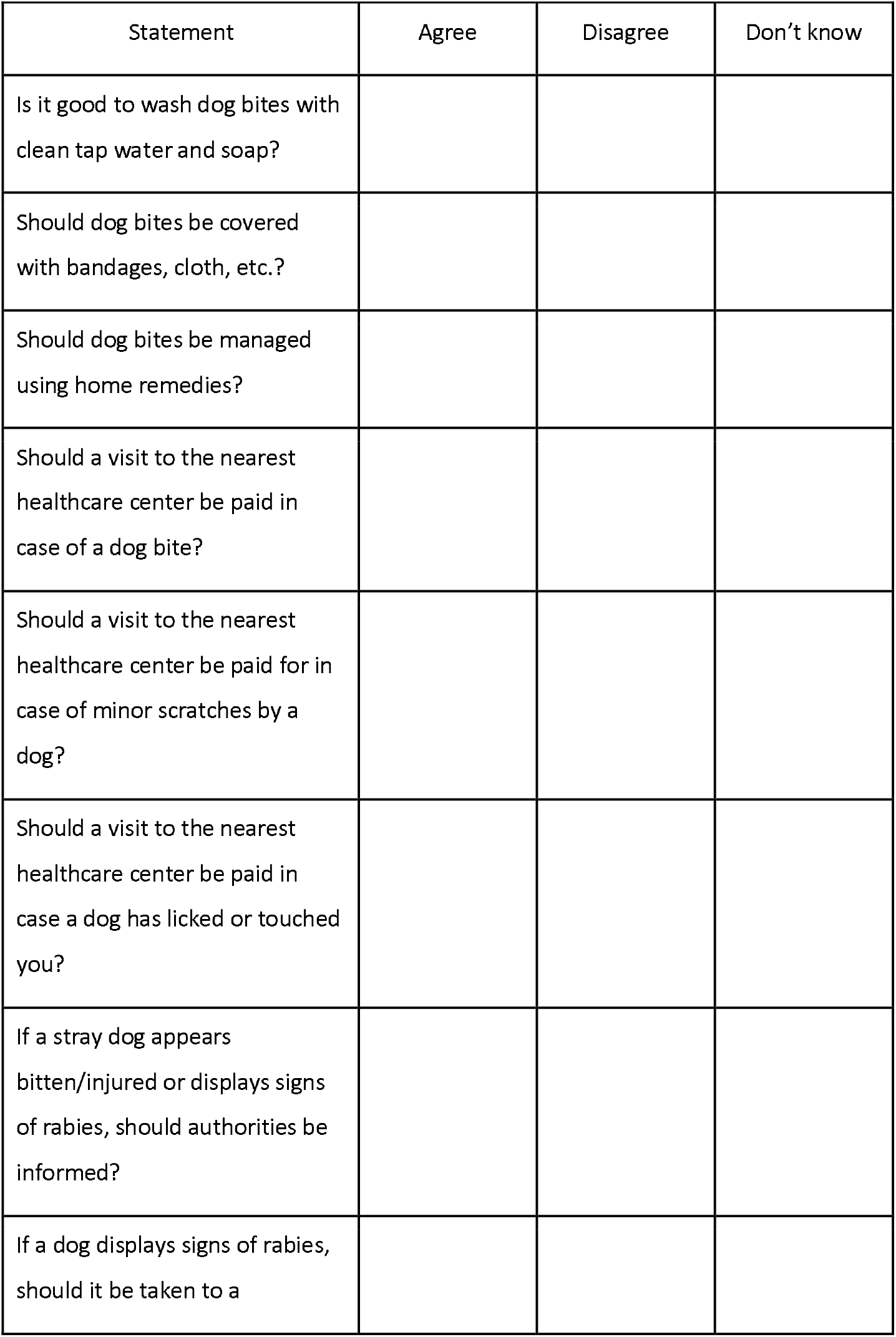
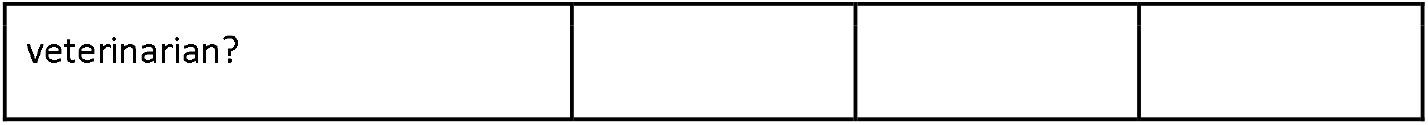

###### Part VI: Changes in IEC Material

Please mention any changes in the IEC material that you would like (text clarity, more pictures, simpler language, etc.).

### Appendix B

Informed Consent Form

#### INFORMED CONSENT FORM

##### Name of the Project

Social Media as an Information, Education, and Communication Tool for Rabies Prevention: An Interventional Study

##### Part I: Information Sheet

###### Introduction

I am Jay Verma, an M.B.B.S. student at Maulana Azad Medical College (MAMC), Delhi. I’m researching to determine the effects of social media on rabies IEC material dissemination and retention. I am going to give you information and invite you to be part of this research. You do not have to decide today whether or not you will participate in the research. Before you decide, you can talk to me about the research. There may be some words that you do not understand. Please ask me to stop as we go through the information and I will take the time to explain. If you have questions later, you can ask them me. It has been approved by the Institutional Ethics Committee of MAMC.

###### Type of Research Intervention

In this study, you will be asked to fill out a questionnaire two times at a gap of nearly 45 days. The questionnaire will take around 20 minutes to fill out. Moreover, you may receive some WhatsApp messages covering the IEC material.

###### Voluntary Participation

It is entirely your choice whether you wish to participate in this study or not. If you decide to participate and then change your mind later on, you are free to opt-out of this study.

###### Confidentiality

All of the information gathered during this research will be kept confidential. During data analysis, your identifying information will be replaced by coded information and no one except the principal investigator will know your identity.

You can ask me any more questions about any part of the research study, if you wish to.

##### Part II: Certificate of Consent

I have read the foregoing information, or it has been read to me. I have had the opportunity to ask questions about it and any questions that I have asked have been answered to my satisfaction. I consent voluntarily to participate as a participant in this research.

Name of Participant

__________________

Signature of Participant

__________________

Date

__________________

## REFERENCES

1. NHP Admin. nhp.gov.in [Internet]. Delhi: NHP Admin; [updated 2018 Dec 18; cited 2022 Feb 3]. Available from: https://www.nhp.gov.in/national-rabies-control-programme_pg

2. Directorate General of Health Services. National Rabies Control Program - Operational Guidelines, 8. [Internet]. 2013 [cited 2022 Feb 3]. Available from: http://www.health.punjab.gov.in/sites/default/files/NRCP%20Operational%20Guidelines.PDF

3. Breland JY, Quintiliani LM, Schneider KL, May CN, Pagoto S. Social media as a tool to increase the impact of public health research. Am J Public Health [Internet]. 2017;107(12):1890–1. Available from: http://dx.doi.org/10.2105/AJPH.2017.304098

4. Cinelli M, Quattrociocchi W, Galeazzi A, Valensise CM, Brugnoli E, Schmidt AL, et al. The COVID-19 social media infodemic. Sci Rep [Internet]. 2020;10(1):16598. Available from: http://dx.doi.org/10.1038/s41598-020-73510-5

5. Dijkstra S, Kok G, Ledford JG, Sandalova E, Stevelink R. Possibilities and pitfalls of social media for translational medicine. Front Med (Lausanne) [Internet]. 2018;5:345. Available from: http://dx.doi.org/10.3389/fmed.2018.00345

6. Gatewood J, Monks SL, Singletary CR, Vidrascu E, Moore JB. Social media in public health: Strategies to distill, package, and disseminate public health research. J Public Health Manag Pract [Internet]. 2019; Available from: http://dx.doi.org/10.1097/PHH.0000000000001096

7. Greene JA, Choudhry NK, Kilabuk E, Shrank WH. Online social networking by patients with diabetes: a qualitative evaluation of communication with Facebook. J Gen Intern Med [Internet]. 2011;26(3):287– 92. Available from: http://dx.doi.org/10.1007/s11606-010-1526-3

8. Liang B, Scammon DL. E-Word-of-Mouth on health social networking sites: An opportunity for tailored health communication: EWOM on health social networking sites. J consum behav [Internet]. 2011;10(6):322–31. Available from: http://dx.doi.org/10.1002/cb.378

9. Rajagopalan MS, Khanna VK, Leiter Y, Stott M, Showalter TN, Dicker AP, et al. Patient-oriented cancer information on the internet: a comparison of wikipedia and a professionally maintained database. J Oncol Pract [Internet]. 2011;7(5):319–23. Available from: http://dx.doi.org/10.1200/JOP.2010.000209

10. Nordqvist C, Hanberger L, Timpka T, Nordfeldt S. Health professionals’ attitudes towards using a Web 2.0 portal for child and adolescent diabetes care: qualitative study. J Med Internet Res [Internet]. 2009;11(2):e12. Available from: http://dx.doi.org/10.2196/jmir.1152

